# Can Large Language Models Aid Caregivers of Pediatric Cancer Patients in Information Seeking? A Cross-Sectional Investigation

**DOI:** 10.1101/2024.08.08.24311711

**Authors:** Emre Sezgin, Daniel I. Jackson, A. Baki Kocaballi, Mindy Bibart, Sue Zupanec, Wendy Landier, Anthony Audino, Mark Ranalli, Micah Skeens

## Abstract

**Background and Objectives:** In pediatric oncology, caregivers seek detailed, accurate, and understandable information about their child’s condition, treatment, and side effects. The primary aim of this study was to assess the performance of four publicly accessible large language model (LLM)- supported knowledge generation and search tools in providing valuable and reliable information to caregivers of children with cancer.

**Methods:** This cross-sectional study evaluated the performance of the four LLM-supported tools — ChatGPT (GPT-4), Google Bard (Gemini Pro), Microsoft Bing Chat, and Google SGE- against a set of frequently asked questions (FAQs) derived from the Children’s Oncology Group Family Handbook and expert input. Five pediatric oncology experts assessed the generated LLM responses using measures including Accuracy (3-point ordinal scale), Clarity (3-point ordinal scale), Inclusivity (3-point ordinal scale), Completeness (Dichotomous nominal scale), Clinical Utility (5-point Likert-scale), and Overall Rating (4-point ordinal scale). Additional Content Quality Criteria such as Readability (ordinal scale; 5- 18th grade of educated reading), Presence of AI Disclosure (Dichotomous scale), Source Credibility (3- point interval scale), Resource Matching (3-point ordinal scale), and Content Originality (ratio scale) were also evaluated. We used descriptive analysis including the mean, standard deviation, median, and interquartile range. We conducted Shapiro-Wilk test for normality, Levene’s test for homogeneity of variances, and Kruskal-Wallis H-Tests and Dunn’s post-hoc tests for pairwise comparisons.

**Results:** Through expert evaluation, ChatGPT showed high performance in accuracy (M=2.71, SD=0.235), clarity (M=2.73, SD=0.271), completeness (M=0.815, SD=0.203), Clinical Utility (M=3.81, SD=0.544), and Overall Rating (M=3.13, SD=0.419). Bard also performed well, especially in accuracy (M=2.56, SD=0.400) and clarity (M=2.54, SD=0.411), while Bing Chat (Accuracy M=2.33, SD=0.456; Clarity M=2.29, SD=0.424) and Google SGE (Accuracy M=2.08, SD=0.552; Clarity M=1.95, SD=0.541) had lower overall scores. The Presence of AI Disclosure was less frequent in ChatGPT (M=0.69, SD=0.46), which affected Clarity (M=2.73, SD=0.266), whereas Bard maintained a balance between AI Disclosure (M=0.92, SD=0.27) and Clarity (M=2.54, SD=0.403). Overall, we observed significant differences between LLM tools (p < .01).

**Conclusions:** LLM-supported tools potentially contribute to caregivers’ knowledge of pediatric oncology on related topics. Each model has unique strengths and areas for improvement, suggesting the need for careful selection and evaluation based on specific clinical contexts. Further research is needed to explore the application of these tools in other medical specialties and patient demographics to assess their broader applicability and long-term impacts, including the usability and feasibility of using LLM-supported tools with caregivers.

## Introduction

Pediatric oncology presents unique challenges not only in treatment but also in the need for accurate information and education provided to caregivers during the course of therapy. Caregivers often seek detailed, accurate, and understandable information regarding their child’s condition, treatment, and side effects ^1^. The need for effective real-time information is critical as it impacts the caregivers’ understanding of cancer treatment and ability to make informed decisions to support their child through the treatment journey. The literature on caregiver information-seeking behaviors reports that a significant population of oncology caregivers rely on informal sources (e.g., the Internet) for health related questions ^2^. An earlier study reported that up to 87% of oncology caregivers are frequently relying on search engines and similar tools due to their convenience and high accessibility ^19^. Traditional information sources (e.g., pamphlets or booklets provided by the clinics), while valuable, may not always provide tailored feedback that caregivers seek during or after clinical visits ^3,4^. This can create a challenging environment where provided resources could be limited to learn about conditions and treatments as well as to find timely answers for unique caregiver concerns. Intelligent systems could bridge the gap and address this challenge by providing just-in-time and tailored responses ^5^.

Artificial Intelligence (AI) models (such as Large Language Models (LLMs)) and their applications have demonstrated notable capabilities in natural language processing tasks, surpassing previous state-of-the-art methods in areas like information retrieval ^6^, question answering ^7^, and medical text summarization ^8^. These tools are increasingly being utilized to improve access to online medical information ^9^. Particularly via publicly available tools (e.g., ChatGPT), health information seeking and access have started to shift from basic keyword-based searching to LLM-based augmented searches. Furthermore, these tools could be valuable in pediatric oncology, providing personalized responses to caregiver queries ^10^. Due to their conversational interface, most of these tools are available to lay users similar to a public search engine (e.g., asking questions to chatGPT without prompt engineering or fine-tuning). This may present a risk for misinformation and misguidance. Therefore, the performance of these AI-driven tools in delivering clinically relevant, accurate, and understandable information needs to be ensured and evaluated. Current literature demonstrates that LLM-based tools can address a wide array of health questions but vary in readability and accuracy based on the domain of healthcare ^11^. To our knowledge, few studies have investigated the AI generated responses from the perspective of pediatric healthcare providers ^12,13^.

The primary aim of this study is to assess AI-supported (more specifically, LLM-supported) tools in providing useful and reliable information to caregivers of children with cancer. Our research question is: “What is the performance of the LLM-supported tools in response to pediatric oncology-related frequently asked questions from patients/caregivers?”

## Methods

In this cross-sectional study, we evaluate the performance of LLM-supported applications. Their responses to frequently asked patient and/or caregiver questions (FAQs) regarding pediatric oncology ^14^ are assessed by a panel of five pediatric oncology experts using a comprehensive set of measures.

### Model Selection

Study team selected the most publicly accessible and generalized LLM-supported applications. These generalized models are preferred because of their availability to public (free to use) and high demand/popularity. Therefore, the selection included two LLM-supported knowledge generation tools [Google Bard/Gemini ^15^, ChatGPT ^16^], and two LLM-supported search tools [Google Search with AI (SGE) ^17^, Bing Chat/Microsoft Copilot ^18^]. From this point forward, we will use “LLM tools” to refer to these 4 platforms throughout this paper.

### Question Creation

The research team members manually created a preliminary set of 10 questions using pediatric oncology topics in the COG Family Handbook ^14^. Each question was phrased to reflect the perspective of a caregiver of a child with cancer. This included rephrasing queries and asking follow-up questions. A board of professional stakeholders (n=3), comprised of COG leadership, reviewed the questions for plausible phrasing and topic relevance. Of the original 10 questions, one was removed as irrelevant, and nine questions were revised. The stakeholders recommended 17 additional questions to enrich the available diagnosis-specific information grouped by disease/treatment phases. Research team combined the suggestions to create a finalized set of 26 frequently asked questions (FAQs) to prompt LLM tools (Table 1).

**Table 1.** Pediatric oncology FAQs grouped by disease/treatment phase.

| **Phases** | **Questions** |
|---|---|
| **Pre-Diagnosis** | 1. What causes childhood cancer?<br>2. Is childhood cancer contagious? |
| **Diagnosis** | 3. How is childhood cancer diagnosed?<br>4. What is a biopsy?<br>5. How will the results of my child’s biopsy be used in planning their treatment?<br>6. What are the symptoms of anemia in a child with cancer? |
| **Cancer Treatment** | 7. What are the treatment options for my child’s cancer?<br>8. Can you specify different treatment options for other types of childhood cancer?<br>9. What should I do if my child has symptoms of anemia?<br>10. What can be done to help my child cope with hair loss?<br>11. Do I need to keep my child with cancer away from other children?<br>12. What causes weight gain during childhood cancer treatment?<br>13. Is there anything I can do to help my child avoid excess weight gain during their treatment?<br>14. What precautions should I take if my child has thrombocytopenia?<br>15. My child with cancer needs a central line. What are the options? |
|  | 16. What are the pros/cons of each type of central line?<br>17. What precautions do I need to take while my child is receiving chemotherapy?<br>18. What should I do if my child gets a fever during cancer treatment?<br>19. Will my child be able to return to school during cancer treatment?<br>20. What precautions are needed if my child returns to school?<br>21. What is a clinical trial?<br>22. What are the pros/cons of enrolling my child on a clinical trial for treatment of their cancer? |
| **Survivorship** | 23. What is the typical follow-up plan once my child completes treatment for cancer?<br>24. How long will my child continue with follow-up after completing cancer treatment?<br>25. What will be done at follow-up visits after completing cancer treatment?<br>26. Are there ways I can help my other children cope with their sibling’s cancer diagnosis? |

### Expert Selection

Five pediatric oncology experts acted as raters (one Nursing Discipline Chair of COG, two oncology nurses with doctoral degrees, and two oncology physicians). Each expert was introduced to the concept of generative AI and its capabilities prior to completing study activities. Experts completed the rating tasks remotely using a REDCap survey to organize their responses.

### Response Generation

Four LLM tools were prompted with the complete set of FAQs in the subject domain of pediatric oncology: ChatGPT [GPT-4 Turbo ^16^], Bard [Gemini Pro patch v2023.12.06 ^15^], Microsoft Bing Chat [renamed as Copilot; supported by ChatGPT-4, Prometheus, and DALL-E 3; set to “More Balanced” mode ^18^], and Google SGE [Generative AI in Search, supported by Gemini Pro, MUM, PaLM 2 ^17^] between 11/30/2023-12/13/2023. Each question was asked in a new session with each LLM tools (not sequentially in a single session). Then, the generated responses were copied to a text editor. Text formatting, including bold, italics, underline, highlighted, and bullet/number lists, was preserved to the greatest degree possible. Images in generative answers were excluded from expert review to standardize results to textual data. Hyperlinks were also removed to reduce bias towards the LLM tools tuned to provide links and ensure uniformity with the generated text only. For questions 12, 13, and 23, Google AI search provided no generated answers, which were excluded from the analysis. To ensure LLM response consistency, the research team prompted each question three times to each LLM tool and reviewed responses within each group (given minimal discrepancy, we selected first iterations throughout the study). Finally, a total of 104 responses were generated from four LLM tools. Responses were recorded in a REDCap form for expert evaluation. An example generated response for a question is given in textbox 1. The list of generated responses can be shared upon request.

Textbox 1. Example generated response by GPT-4 via ChatGPT (prompted on 12/2023)

**Question:** How will the results of my child’s biopsy be used in planning their treatment?

**Response:** The results of your child’s biopsy play a crucial role in planning their treatment. Here’s how the process typically unfolds:

1. *Diagnosis Confirmation: The primary purpose of a biopsy is to confirm a diagnosis. Depending on what the doctors suspect (like a tumor, infection, or other condition), the biopsy will reveal the exact nature of the issue*.
2. *Determining the Type and Stage: Especially in the case of cancer, a biopsy can determine not just the presence of cancerous cells, but also the specific type and stage of the cancer. This information is vital for deciding the most effective treatment approach*.
3. *Tailoring Treatment Plans: Treatment plans are often customized based on the biopsy results. For instance, certain types of cancer respond better to certain medications, and the choice of chemotherapy or radiation therapy, if needed, often depends on the biopsy findings*.
4. *Identifying Genetic Markers: Some biopsies can reveal genetic markers that are significant for treatment. In cancer, for example, certain genetic mutations might make the cancer more susceptible to specific drugs*.
5. *Monitoring Response to Treatment: In some cases, repeat biopsies are performed during or after treatment to see how the disease is responding. This helps in adjusting treatment plans if necessary*.
6. *Guiding Surgical Decisions: If surgery is a treatment option, the biopsy results can help in planning the extent of surgery required. This is particularly true in cancer treatment, where the goal is often to remove all cancerous tissue*.

*Remember, the exact use of biopsy results can vary depending on the specific medical condition and individual circumstances. It’s important to discuss with your child’s healthcare team how the biopsy results will inform their particular treatment plan*.

### Evaluation Criteria and Measures

We created a set of evaluation criteria and associated measures to assess the performance of LLM tools. Two groups of measures were created. (1) Expert Evaluation Criteria include Accuracy, Clarity, Inclusivity, Completeness, Clinical Utility, and Overall Rating (Table 2 for descriptions and scales). These measures were adapted from literature investigating clinical accuracy among LLM tools ^20^. Additionally, experts provided free-text response to share feedback about responses and their ratings. Experts were provided the measures in the REDCap form. (2) Content Quality Criteria include Readability, Presence of AI Disclosure, Source Credibility, Resource Matching, and Content Originality ^10,21–23^. Content quality analysis was conducted by the research team. Table 3 outlines the measures, descriptions, and sources.

**Table 2.**
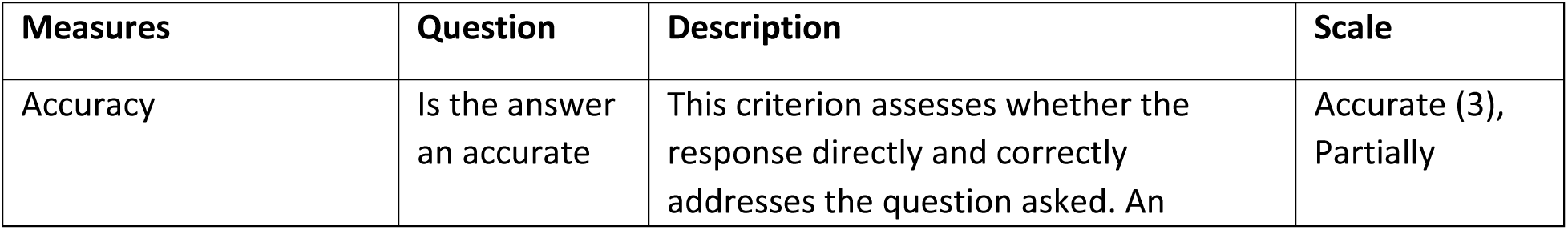
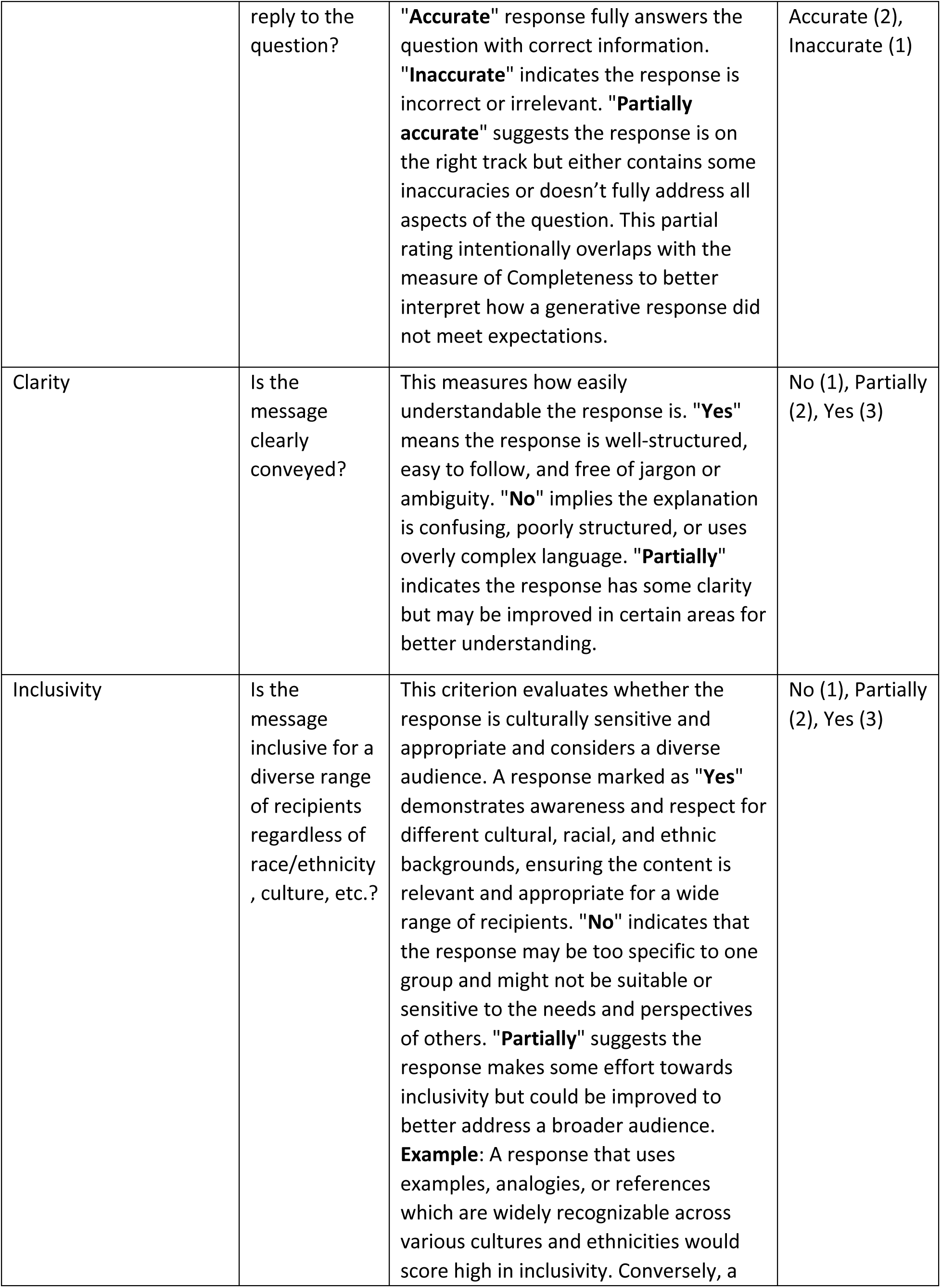
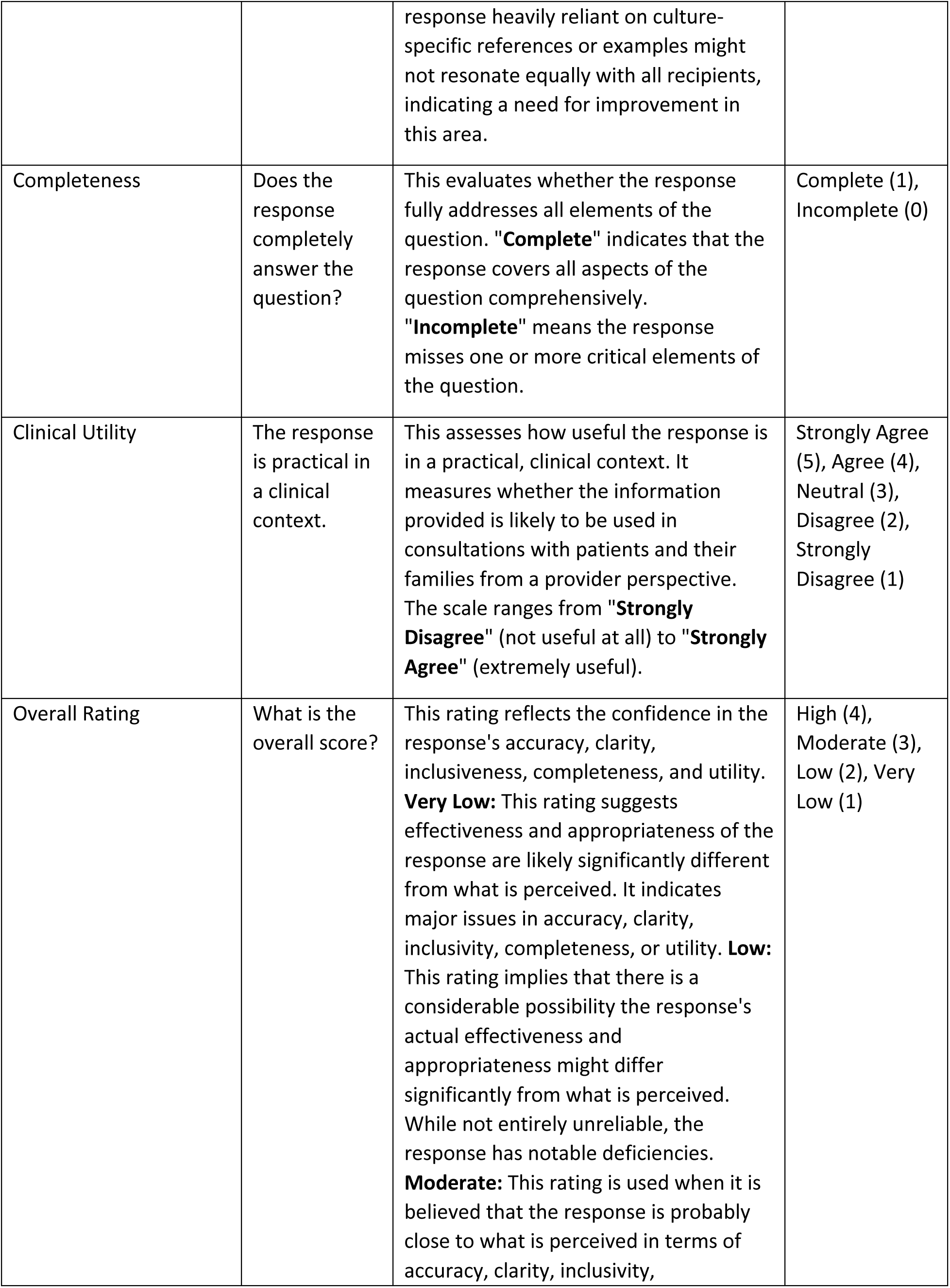
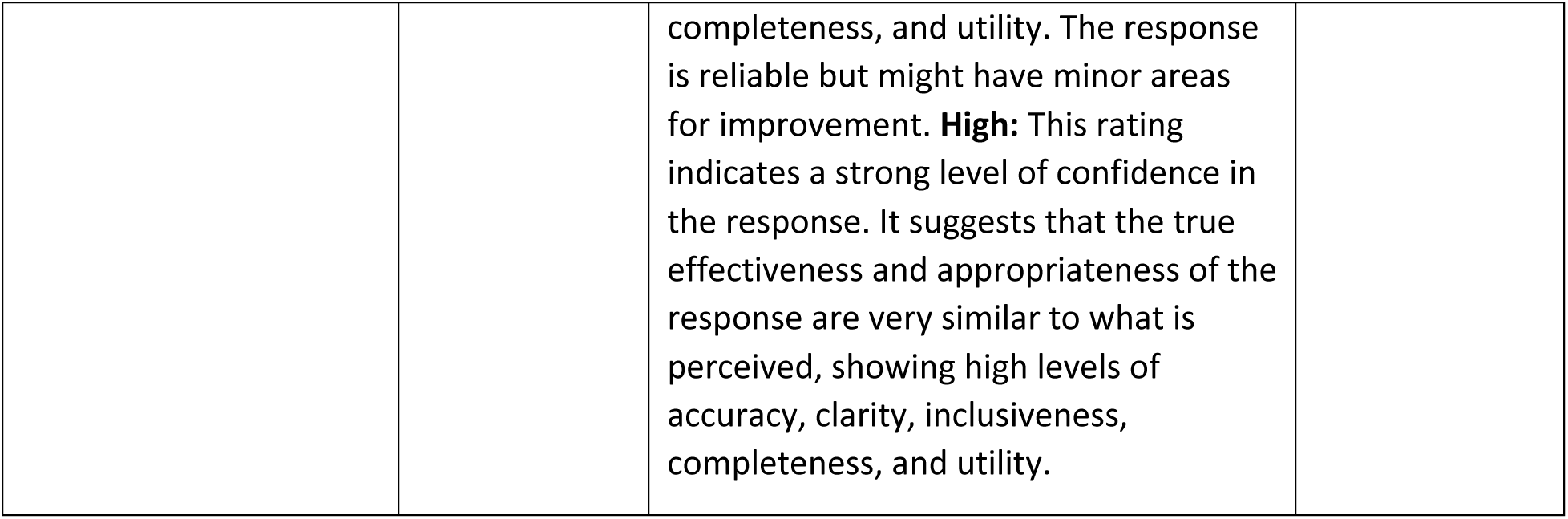
Expert Evaluation Criteria measures and descriptions.

| Measures | Question | Description | Scale |
|---|---|---|---|
| Accuracy | Is the answer an accurate reply to the question? | This criterion assesses whether the response directly and correctly addresses the question asked. An "**Accurate**" response fully answers the question with correct information. "**Inaccurate**" indicates the response is incorrect or irrelevant. "**Partially accurate**" suggests the response is on the right track but either contains some inaccuracies or doesn’t fully address all aspects of the question. This partial rating intentionally overlaps with the measure of Completeness to better interpret how a generative response did not meet expectations. | Accurate (3), Partially Accurate (2), Inaccurate (1) |
| Clarity | Is the message clearly conveyed? | This measures how easily understandable the response is. "**Yes**" means the response is well-structured, easy to follow, and free of jargon or ambiguity. "**No**" implies the explanation is confusing, poorly structured, or uses overly complex language. "**Partially**" indicates the response has some clarity but may be improved in certain areas for better understanding. | No (1), Partially (2), Yes (3) |
| Inclusivity | Is the message inclusive for a diverse range of recipients regardless of race/ethnicity, culture, etc.? | This criterion evaluates whether the response is culturally sensitive and appropriate and considers a diverse audience. A response marked as "**Yes**" demonstrates awareness and respect for different cultural, racial, and ethnic backgrounds, ensuring the content is relevant and appropriate for a wide range of recipients. "**No**" indicates that the response may be too specific to one group and might not be suitable or sensitive to the needs and perspectives of others. "**Partially**" suggests the response makes some effort towards inclusivity but could be improved to better address a broader audience. **Example**: A response that uses examples, analogies, or references which are widely recognizable across various cultures and ethnicities would score high in inclusivity. Conversely, a response heavily reliant on culture-specific references or examples might not resonate equally with all recipients, indicating a need for improvement in this area. | No (1), Partially (2), Yes (3) |
| Completeness | Does the response completely answer the question? | This evaluates whether the response fully addresses all elements of the question. "**Complete**" indicates that the response covers all aspects of the question comprehensively. "**Incomplete**" means the response misses one or more critical elements of the question. | Complete (1), Incomplete (0) |
| Clinical Utility | The response is practical in a clinical context. | This assesses how useful the response is in a practical, clinical context. It measures whether the information provided is likely to be used in consultations with patients and their families from a provider perspective. The scale ranges from "**Strongly Disagree**" (not useful at all) to "**Strongly Agree**" (extremely useful). | Strongly Agree (5), Agree (4), Neutral (3), Disagree (2), Strongly Disagree (1) |
| Overall Rating | What is the overall score? | This rating reflects the confidence in the response's accuracy, clarity, inclusiveness, completeness, and utility. **Very Low:** This rating suggests effectiveness and appropriateness of the response are likely significantly different from what is perceived. It indicates major issues in accuracy, clarity, inclusivity, completeness, or utility. **Low:** This rating implies that there is a considerable possibility the response's actual effectiveness and appropriateness might differ significantly from what is perceived. While not entirely unreliable, the response has notable deficiencies. **Moderate:** This rating is used when it is believed that the response is probably close to what is perceived in terms of accuracy, clarity, inclusivity, completeness, and utility. The response is reliable but might have minor areas for improvement. **High:** This rating indicates a strong level of confidence in the response. It suggests that the true effectiveness and appropriateness of the response are very similar to what is perceived, showing high levels of accuracy, clarity, inclusiveness, completeness, and utility. | High (4), Moderate (3), Low (2), Very Low (1) |

**Table 3.** Content Quality Criteria and descriptions.

| Measure | Description | Scale | References |
|---|---|---|---|
| Readability | An online health literacy editor (SHELL) was used to compute the ease of understanding of texts. This includes the Grade reading score as well as uncommon words, public health jargon, passive voice, and the use of acronyms. | 5-18th (grade of educated reading), Missing (NA) | (Ayre et al., 2023) |
| Presence of AI Disclosure | A measure of whether the generative response contains reminders for the reader to not anthropomorphize the tool, recognize the risk of misinformation, or address a medical professional for more information. | Yes (1), No (0), Not Sure (NA) | n/a |
| Source Credibility | A measure of whether linked resources in a generative answer are recent (Currency), created by experts in the field (Authorship), and published transparently (Attribution) with possible conflicts of interest (Disclosure). Each of the 4 properties contributes 1 out of 4 possible points. | Not Credible (0), Partially Credible (1-3), Credible (4), Missing (NA) | (Hristidis et al., 2023; O’Hagan et al., 2023) |
| Resource Matching | Does the linked resource support the topic described in the generative response? | Full (3), Partial (2), None (1), Missing (NA) | (Liu et al., 2023; Menick et al., 2022) |
| Content Originality | A measure of total words per phrase longer than 5 words in a generative answer identified by Turnitin.com to derive from another online source. | Percentage of verbatim phrases (0-100%) | n/a |

### Statistical Analysis

We reported descriptive analysis including the mean scores, standard deviation, median, and interquartile range. The Cohen Kappa and Weighted Kappa were calculated for pairs of experts. We anticipated that occupational roles may affect interrater agreement, therefore experts were grouped into 2 groups to calculate paired agreement: Nurses and Physicians. In addition, statistical analyses were conducted to explore the differences between LLM tools. Expert evaluation criteria were tested for normality using the Shapiro-Wilk method and the Kruskal-Wallis H-Test to report significant differences. Then we calculated the variance of homogeneity using the Levene Test. Pairs of LLM tools were calculated with the Dunn Test for measuring similarity between models in a matrix. IBM Statistical Package for the Social Sciences (SPSS) Statistics on Version: 28.0.0.0 (190) and RStudio 2023.12.1 Build 402 were used for analysis and visuals ^24,25^.

## Results

Under the expert evaluation, we reported the performance of four LLM tools across six measures that were rated by five pediatric oncology experts: Accuracy, Clarity, Inclusivity, Completeness, Clinical Utility, and Overall Rating (Table 4, Figure 1). Raters demonstrated moderate agreement during the assessment (Average Cohen’s k=0.65).

**Figure 1.**
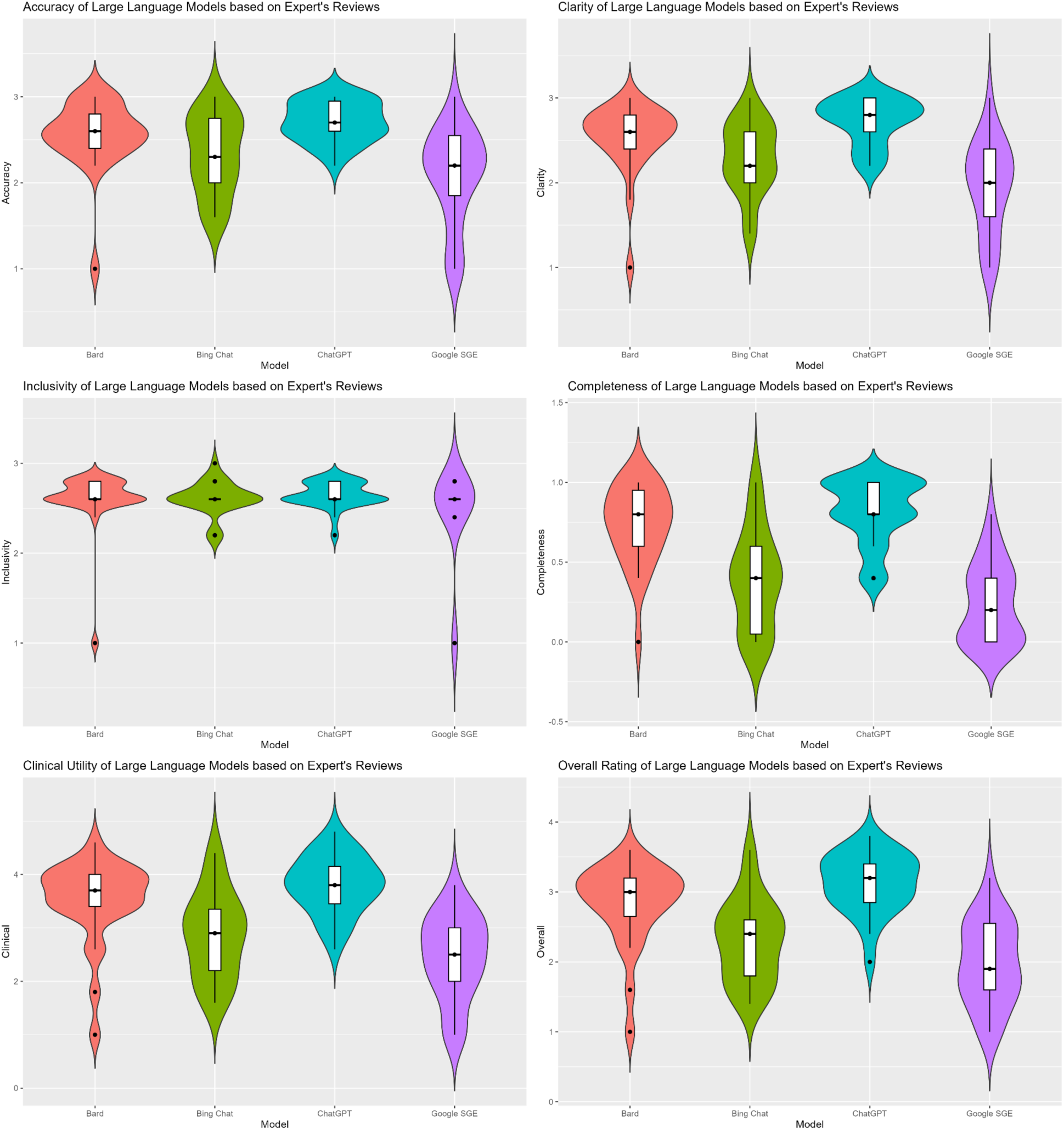
Violin graphs of Expert Evaluation Criteria

**Table 4.**
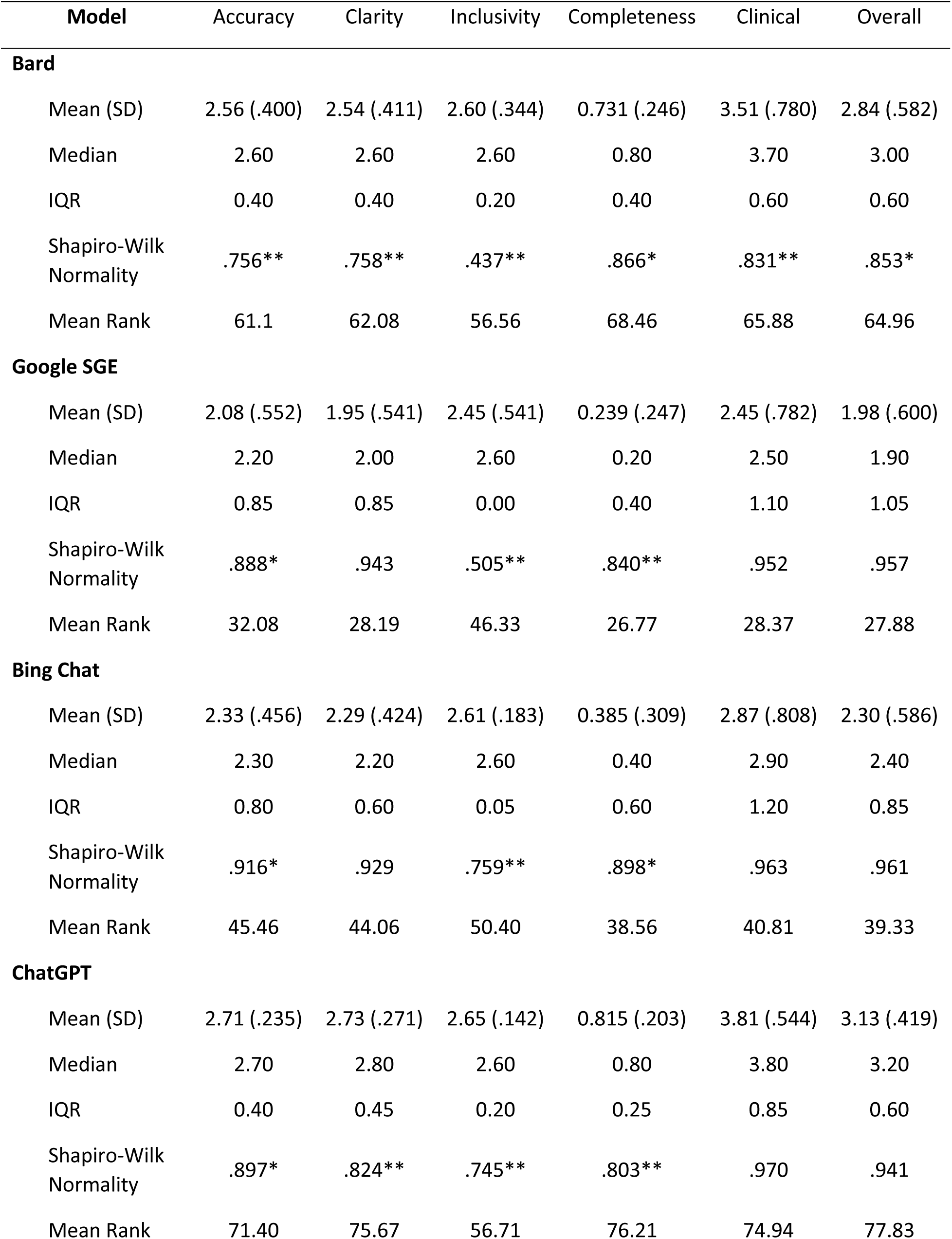
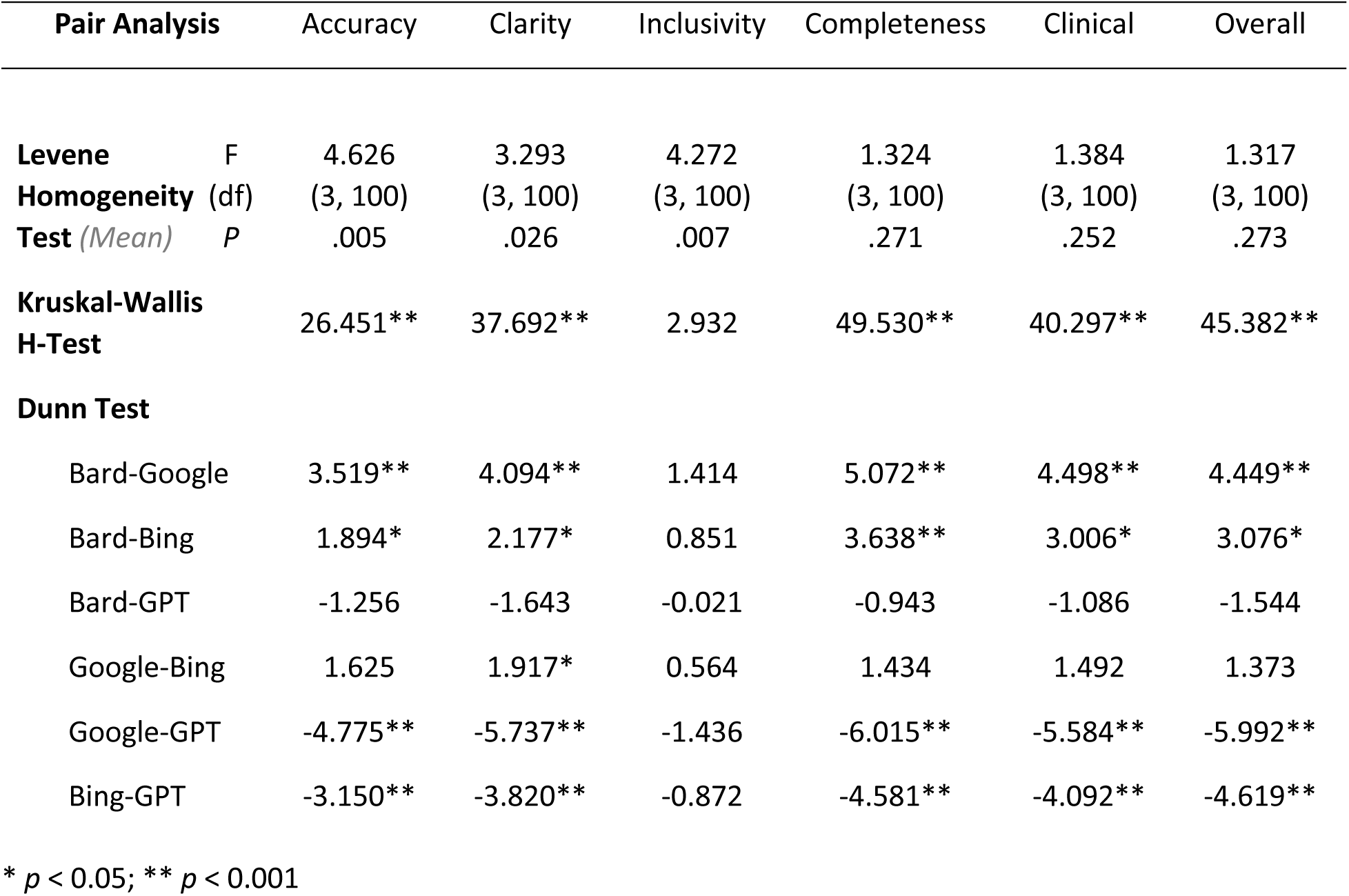
LLM tools organized by Expert Evaluation Criteria from 5 experts.

| **Model** | Accuracy | Clarity | Inclusivity | Completeness | Clinical | Overall |
|---|---|---|---|---|---|---|
| **Bard** |  |  |  |  |  |  |
| Mean (SD) | 2.56 (.400) | 2.54 (.411) | 2.60 (.344) | 0.731 (.246) | 3.51 (.780) | 2.84 (.582) |
| Median | 2.60 | 2.60 | 2.60 | 0.80 | 3.70 | 3.00 |
| IQR | 0.40 | 0.40 | 0.20 | 0.40 | 0.60 | 0.60 |
| Shapiro-Wilk Normality | .756** | .758** | .437** | .866* | .831** | .853* |
| Mean Rank | 61.1 | 62.08 | 56.56 | 68.46 | 65.88 | 64.96 |
| **Google SGE** |  |  |  |  |  |  |
| Mean (SD) | 2.08 (.552) | 1.95 (.541) | 2.45 (.541) | 0.239 (.247) | 2.45 (.782) | 1.98 (.600) |
| Median | 2.20 | 2.00 | 2.60 | 0.20 | 2.50 | 1.90 |
| IQR | 0.85 | 0.85 | 0.00 | 0.40 | 1.10 | 1.05 |
| Shapiro-Wilk Normality | .888* | .943 | .505** | .840** | .952 | .957 |
| Mean Rank | 32.08 | 28.19 | 46.33 | 26.77 | 28.37 | 27.88 |
| **Bing Chat** |  |  |  |  |  |  |
| Mean (SD) | 2.33 (.456) | 2.29 (.424) | 2.61 (.183) | 0.385 (.309) | 2.87 (.808) | 2.30 (.586) |
| Median | 2.30 | 2.20 | 2.60 | 0.40 | 2.90 | 2.40 |
| IQR | 0.80 | 0.60 | 0.05 | 0.60 | 1.20 | 0.85 |
| Shapiro-Wilk Normality | .916* | .929 | .759** | .898* | .963 | .961 |
| Mean Rank | 45.46 | 44.06 | 50.40 | 38.56 | 40.81 | 39.33 |
| **ChatGPT** |  |  |  |  |  |  |
| Mean (SD) | 2.71 (.235) | 2.73 (.271) | 2.65 (.142) | 0.815 (.203) | 3.81 (.544) | 3.13 (.419) |
| Median | 2.70 | 2.80 | 2.60 | 0.80 | 3.80 | 3.20 |
| IQR | 0.40 | 0.45 | 0.20 | 0.25 | 0.85 | 0.60 |
| Shapiro-Wilk Normality | .897* | .824** | .745** | .803** | .970 | .941 |
| Mean Rank | 71.40 | 75.67 | 56.71 | 76.21 | 74.94 | 77.83 |
| **Pair Analysis** | Accuracy | Clarity | Inclusivity | Completeness | Clinical | Overall |
| **Levene Homogeneity Test** *(Mean)* |  |  |  |  |  |  |
| F | 4.626 | 3.293 | 4.272 | 1.324 | 1.384 | 1.317 |
| (df) | (3, 100) | (3, 100) | (3, 100) | (3, 100) | (3, 100) | (3, 100) |
| *P* | .005 | .026 | .007 | .271 | .252 | .273 |
| **Kruskal-Wallis H-Test** | 26.451** | 37.692** | 2.932 | 49.530** | 40.297** | 45.382** |
| **Dunn Test** |  |  |  |  |  |  |
| Bard-Google | 3.519** | 4.094** | 1.414 | 5.072** | 4.498** | 4.449** |
| Bard-Bing | 1.894* | 2.177* | 0.851 | 3.638** | 3.006* | 3.076* |
| Bard-GPT | -1.256 | -1.643 | -0.021 | -0.943 | -1.086 | -1.544 |
| Google-Bing | 1.625 | 1.917* | 0.564 | 1.434 | 1.492 | 1.373 |
| Google-GPT | -4.775** | -5.737** | -1.436 | -6.015** | -5.584** | -5.992** |
| Bing-GPT | -3.150** | -3.820** | -0.872 | -4.581** | -4.092** | -4.619** |

\* *p* < 0.05; \*\* *p* < 0.001

In terms of Accuracy, ChatGPT had the highest mean score (M=2.71, SD=0.235), followed by Bard (M=2.56, SD=0.400), Bing Chat (M=2.33, SD=0.456), and Google SGE (M=2.08, SD=0.552). The Kruskal-Wallis H-test confirmed significant differences in accuracy among the models (H=26.451, p<.001). Dunn’s test indicated that ChatGPT was significantly more accurate than both Google SGE (p<.001) and Bing Chat (p<.01). Clarity scores were highest for ChatGPT (M=2.73, SD=0.271), with following Bard (M=2.54, SD=0.411), Bing Chat (M=2.29, SD=0.424), and Google SGE (M=1.95, SD=0.541). The differences in clarity were statistically significant (H=37.692, p<.001). Dunn’s test showed significant clarity differences between ChatGPT and both Google SGE (p<.001) and Bing Chat (p<.01). Inclusivity scores were relatively similar across the models, with ChatGPT scoring highest (M=2.65, SD=0.142), then Bing Chat (M=2.61, SD=0.183), Bard (M=2.60, SD=0.344), and Google SGE (M=2.45, SD=0.541). The Kruskal-Wallis H-test indicated no significant differences in inclusivity (H=2.932, p=.403). Completeness scores were highest for ChatGPT (M=0.815, SD=0.203), followed by Bard (M=0.731, SD=0.246), Bing Chat (M=0.385, SD=0.309), and Google SGE (M=0.239, SD=0.247). Significant differences were found in completeness (H=49.530, p<.001). Dunn’s test revealed significant differences between ChatGPT and both Google SGE (p<.001) and Bing Chat (p<.001). Clinical Utility scores were highest for ChatGPT (M=3.81, SD=0.544), with following Bard (M=scoring 3.51, SD=0.780), Bing Chat (M=2.87, SD=0.808) and Google SGE (M=2.45, SD=0.782). The Kruskal-Wallis H-test showed significant differences in Clinical Utility (H=40.297, p<.001). Dunn’s test found ChatGPT significantly more clinically relevant than both Google SGE (p<.001) and Bing Chat (p<.01). Overall Rating scores were highest for ChatGPT (M=3.13, SD=0.419), followed by Bard (M=2.84, SD=0.582), Bing Chat (M=2.30, SD=0.586), and Google SGE (M=1.98, SD=0.600). The differences in Overall Rating were significant (H=45.382, p<.001). Dunn’s test showed ChatGPT had a significantly higher Overall Rating than both Google SGE (p<.001) and Bing Chat (p<.001).

Content Quality results presented notable differences in LLM tool performance on AI Disclosure, Source Credibility, and Resource Matching (Figure 2). AI Disclosure has been chiefly adhered to by Bard (M=0.92, SD=0.27), less by Bing Chat (M=0.62, SD=0.49) and ChatGPT (M=0.69, SD=0.46). Source Credibility was high with Bing Chat (M=2.7, SD=0.61) and Google SGE (M=2.8, SD=0.51), whereas minimally by ChatGPT (M=1.2, SD=0.53). Similarly, resource matching was highly scored for Bing Chat (M=2.7, SD=0.53) and Google SGE (M=2.8, SD=0.51), and minimally by ChatGPT (M=1.2, SD=0.53).

**Figure 2.**
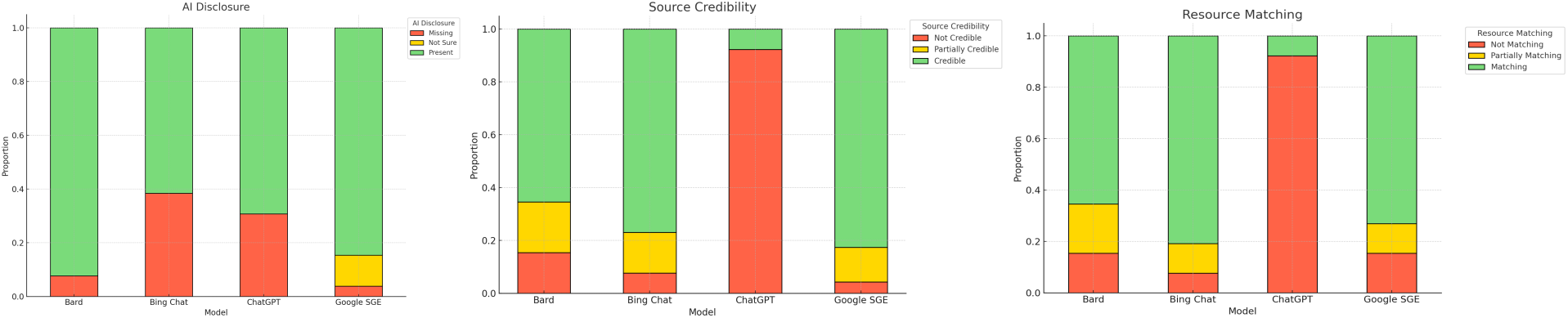
AI Disclosure, Source Credibility, and Resource Matching stacked bar graphs

ChatGPT and Bard have relatively high likelihood of producing original text (with 14-15% matching content with online resources). In contrast, Bing Chat showed greater tendency to produce content that closely matches existing materials online, (M=0.58, SD=0.18) similar to Google SGE.

The models show variability in their Readability scores (Figure 3). However, all LLM tools scored higher than 8th grade readability (which is expected for health information). ChatGPT often scores higher (>13^th^ grade), providing more complex responses. The Bard model tends to generate more extensive responses with more characters and words and a tendency to provide more detailed answers. On the other hand, Google SGE occasionally produces shorter responses, using lower character and word counts, possibly reflecting a more concise answering style. Bing Chat’s performance is mixed, with its readability scores sometimes on the lower end, hinting at simpler or less verbose responses. Across all models, there is a variation in the use of unique words, sentences, and paragraphs, highlighting differences in style and substance in the answers provided (Bard M=177.1, SD=43; Bing Chat M=108.6, SD=29.1; ChatGPT M=159.7, SD=34.2; Google SGE M=92.2, SD=16.9).

**Figure 3.**
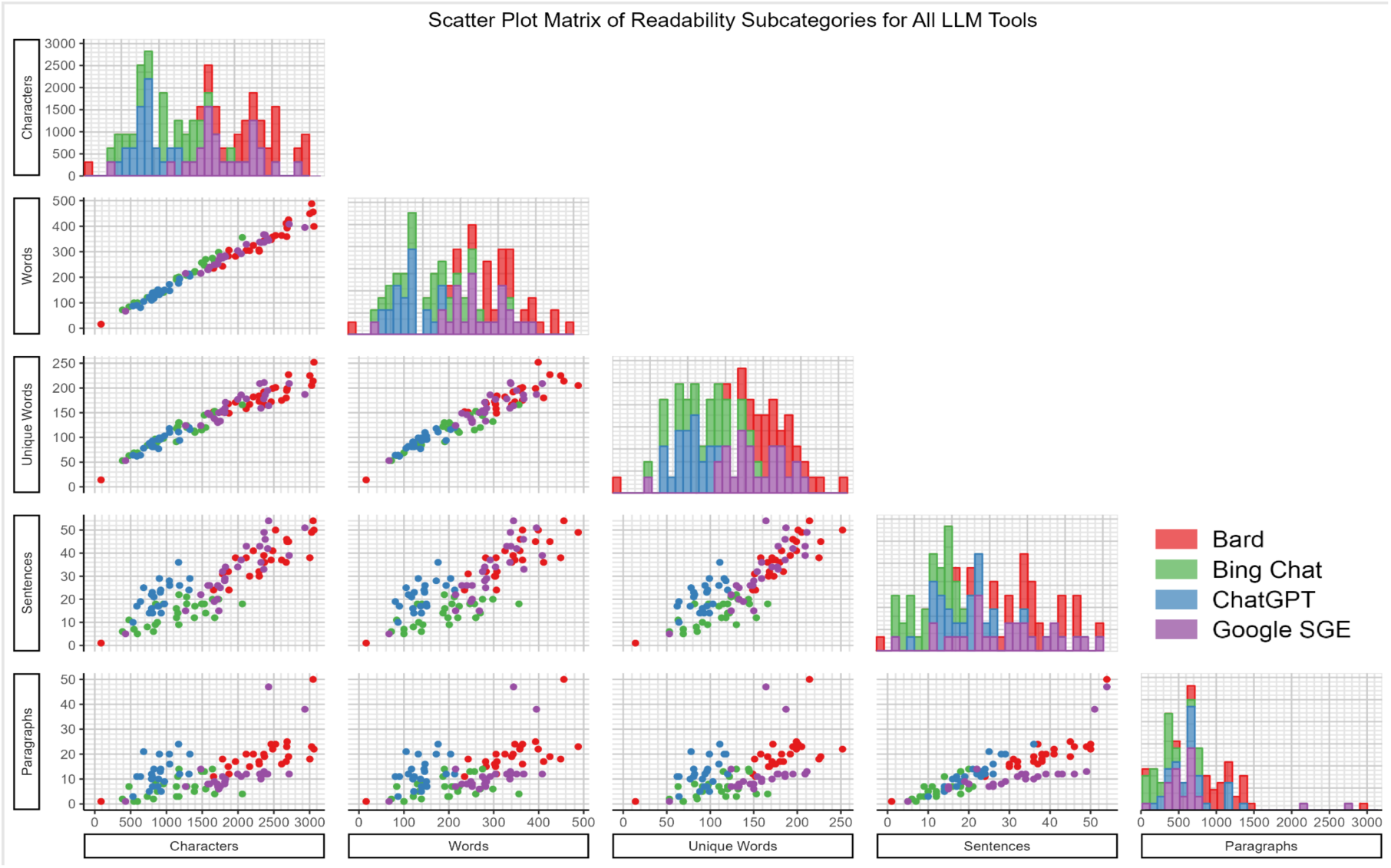
Readability scatterplot matrix

### Qualitative Analysis of Expert Evaluation Criteria Notes

The research team reviewed the free-text notes associated with expert ratings reported for each of the LLM responses. In total, 271 free-text responses were reviewed and analyzed to identify differentiating and common themes (See Appendix 1).

### Experts’ Comments on LLM Tools

We found that Google SGE received criticism for using complex language and inaccuracies, such as the misleading claim that cancer cannot be inherited and incorrect generalizations about cancer patients’ school attendance. Some responses were vague or incomplete, lacking practical advice or clear instructions. Emotional support was sometimes inadequate, with a lack of empathy and an overly technical tone noted by some users.

Bard, another Google LLC product, was also critiqued for using overly complex language, use of technical terms, inaccuracies in details and a lack of essential information (e.g., in response to “What should I do if my child has symptoms of anemia?”, it provided inaccurate or limited information on the likelihood of hair regrowth). Feedback varied on Bard’s thoroughness, with some raters finding it overly detailed or incomplete. Practical guidance was often not clear, and the empathetic tone received mixed reactions, being appreciated by some but considered overly personal or inappropriate by others.

Bing Chat’s responses were critiqued for using a high reading level, making them difficult for patients and families to understand. Issues included inaccuracies, irrelevant information, and inappropriate examples, such as mentioning "prostate cancer" in pediatric oncology contexts. Important details were often missing, reducing the responses’ usefulness. Practical suggestions were sometimes unrealistic or irrelevant. The empathetic tone was occasionally seen as unnecessary or misdirected, and some responses were criticized for sounding punitive.

ChatGPT’s responses were generally clear and well-organized, praised for their thoroughness and readability. However, advanced language and complex explanations were reported as barriers for non-medical audiences. Accuracy varied, with some responses being comprehensive while others had critical gaps or misleading details. Practical advice was appreciated when provided, though some responses were too generic or not tailored to specific situations. Empathy was positively received but lacked cultural considerations, and some empathetic statements were occasionally inappropriate.

### Qualitative Themes

We identified five common themes across expert feedback regarding LLM tools, which are (1) accessibility of language, (2) accuracy, (3) completeness, (4) practical guidance, and (5) Emotional tone and empathy. Across all models, the *accessibility of language* remains a significant challenge. Despite the advanced capabilities, the responses were found to employ complex medical jargon, posing comprehension difficulties for laypersons. This observed highly with Bard and Bing Chat, while Google SGE and ChatGPT performed relatively better but still inadequate to provide responses in simplified language. *Accuracy* varied notably among the LLMs. All models were prone to inaccuracies, such as Google SGE and Bing Chat were occasionally observed to share misleading medical information. Bard and ChatGPT also displayed inaccuracies, but their responses usually received praise for being comprehensive and clear.

In terms of *completeness*, all LLMs showed mixed results. While Bard and ChatGPT were often recognized for their thoroughness, they sometimes missed critical details or provided excessive, overwhelming information. On the other hand, Google SGE and Bing Chat were often criticized for vague and incomplete responses, suggesting a need for a more balanced and comprehensive approach to information delivery. *Practical guidance* was another area of concern. Across the board, LLMs frequently failed to provide actionable and relevant advice. Google SGE’s responses were noted for their vagueness, and Bing Chat’s practical relevance was often questioned. Bard and ChatGPT provided some practical suggestions, but these were not consistently applicable or tailored to specific user contexts, indicating a gap in practical utility. *Emotional tone* and empathy in responses received mixed feedback. Raters appreciated attempts at empathetic communication, particularly in ChatGPT’s responses, but there were instances where the tone was either too personal or inappropriate, as seen in Bard and Bing Chat.

## Discussion

This study comprehensively evaluates four LLM tools—ChatGPT, Bard, Bing Chat, and Google SGE—in generating responses to frequently asked questions in pediatric oncology. Our expert analysis highlights significant differences in performance across several critical metrics: Accuracy, Clarity, Completeness, Clinical Utility, and Overall Rating.

ChatGPT achieved the highest mean accuracy score, followed by Bard, Bing Chat, and Google SGE, respectively. The statistical analysis revealed significant differences in accuracy, with ChatGPT substantially outperforming Google SGE and Bing Chat. This aligns with prior evidence on search behaviors where ChatGPT outperforms generative search engines when answering FAQs ^26^. ChatGPT provides more precise and correct information, which is crucial for caregivers seeking reliable data and for contributing to online health education ^27^. Similar to accuracy, ChatGPT scored the highest in clarity and completeness, followed by Bard, while Bing Chat and Google SGE showed lower performance. This could result from how these LLM tools are tuned (as ChatGPT and Bard are more generalized models, whereas Google SGE and Bing Chat are more search oriented). However, compromising clarity to benefit quicker web search summaries may not contribute to the need for understandable and clear feedback for caregiver questions. Yet, the nature of rapid summaries as a design philosophy may be a product of current caregiver interactions with popular search tools ^28–30^. The faster, less reliable responses inherently pose a greater risk of error due to lack clinical understanding, but they may also satisfy the target audiences’ expectations despite lower quality and partial responses. Although the focus of the FAQs is aligned with how pediatric oncology caregivers ask about the healthcare journey, personal preferences matter. A caregiver who is looking for quick feedback instead of in-depth information would likely benefit the greatest by alternating between LLM tools rather than relying on one service over the rest. This pattern already exists in the online audience, where caregivers will not only cross-verify online healthcare advice between personal connections and support groups ^31^. An alternative approach may involve an expandable response structure where a concise response can be expanded to provide further details and links for the relevant resources.

An important finding was that LLM tools were, overall, able to score above average with clinically relevant responses. More generalist models (ChatGPT and Bard) had the highest Clinical Utility score, compared to Bing Chat and Google SGE. This was reflected in the raters’ perceived Overall Rating scores as well. This finding aligns with prior literature on LLM performance in the context of clinical knowledge^32^. However, significant differences in Clinical Utility underscore how generalized models outperform search-oriented models. We suspect that LLM tools tuned for generalist knowledge can generate better responses compared to those tuned to web search and summarization for niche and specialized topics like pediatric oncology. However, the margin of error is much narrower for healthcare, let alone pediatrics. Therefore, LLM tools with improved safeguards against misinformation on the internet are likely the models more suited to serve caregivers ^33^.

We found that all models were equally sensitive to cultural diversity without skewing their responses and were universally applicable to a diverse audience of caregivers. Yet, more in-depth research is needed to understand the extent of biases introduced in the LLM tools via alternative testing approaches, such as adversarial testing. This will ensure the potential of LLM tools could be primed to cause misguidance or misinformation ^34–36^.

As discussed in the literature, LLM tools can help make complex information easier to understand and provide real-time personalized education ^37^. Our study found that the readability of responses from different LLM tools varied. ChatGPT often produced well-structured, complex responses with high readability scores, suitable for more educated caregivers, but challenging for those with lower health literacy ^38^. Bard generated even longer, more detailed responses, increasing unique word counts and depth but at the expense of readability. Augmented search tools like Bing Chat and Google SGE provided mixed results, sometimes offering simpler, more straightforward responses that were easier to understand but might lack detail. These overarching trends play a significant role in how caregivers should approach LLM tools in the future, as different platforms in our study demonstrated unique strengths and weaknesses. Unfortunately, all LLM tools scored far from the target 8th grade readability standard for online health information, which is most likely a larger limitation of the technology behind LLM tools ^39^.

### Transparency and Reliability of LLM Tools

AI Disclosure is essential to mitigate the risk of misinformation and increase transparency. This has been adopted by recent clinical tools as well, aiming to notify users that the information received is generated with AI assistance, and professional consultation might be necessary for the vital information received from these LLM tools ^40^. We found that Bard consistently used AI Disclosures, while ChatGPT rarely included in-text AI Disclosures. Augmented search tools moderately adhered to AI Disclosures, adversely impacting transparency and clarity for users’ understanding of the origin of information. Other LLM tools in this study (including ChatGPT) have implemented generic warnings and disclosures as either pop-up notifications or fine prints. However, these additions should be more visible and clearly communicated.

The reliability of the responses was affected by their credibility, matching with resources shared, and content originality. Bard and Bing Chat frequently linked to recent, expert-created resources to enhance trustworthiness. Google SGE performed moderately well, offering credible sources but with less consistency than Bard and Bing Chat. Similarly, aligning response content with supporting materials is essential for reliability. Bard and Bing Chat consistently linked to relevant and supportive resources, ensuring comprehensive information. Google SGE exhibited moderate performance, with occasional gaps in alignment. However, ChatGPT had lower credibility scores due to its design, which is tuned to not share resources unless explicitly prompted to, inherently affecting its performance with our metrics. In line with that, ChatGPT showed weaker performance in resource matching, again influenced by its design to avoid sharing resources, leading to a lack of alignment between content and references. Yet, ChatGPT and Bard demonstrated high Content Originality with low dependency on the exact syntax of the linked resources. Bing Chat, however, had a higher tendency to produce content that closely matched existing materials, followed by Google SGE. Based on the performance of these LLM tools, Content Originality did not necessarily equate to be reliable information (as the content source may vary). As caregivers look for information from credible resources rather than original thoughts, it might be more beneficial to use content from credible resources as-is to convey the message as intended by the professionals. However, this also requires LLMs to ensure using reliable sources while curating responses. In the future, multi-agent approaches might be necessary to assign new LLM roles as the cross-validator of content curated with reliable resources ^41–43^.

### Implications for Personalized Patient/Caregiver Education and Clinical Practices

The application of LLMs in real-time personalized patient information sharing and education suggests a transformative shift in how caregivers’ access and utilize medical information. Our findings indicate that LLMs may enhance the accessibility of complex medical information for non-experts. This is particularly crucial in pediatric oncology, where understanding the nuances of care and treatment options impacts decision-making processes for caregivers ^44,45^. Real-time access to personalized information could bridge gaps in understanding and empower caregivers, leading to more informed discussions with healthcare providers. The ability of LLMs to provide comprehensive and accessible explanations can support more informed decision-making by patients and caregivers. This is especially important in oncology, where treatment decisions can be complex and involve weighing multiple factors ^46^. Real-time, personalized education ensures caregivers know all potential options and the associated risks and benefits. Although it is possible to provide accurate, clear, and clinically useful information, the LLM-based applications in general are not usually consistent to do so (e.g., potential for fabricating facts or hallucinations), which raises concern regarding how lay-user would be able to assess accuracy of information. Thus, use of such applications in pediatric oncology for patient/family information sharing or as educational tool is not considered to be ready for implementation. Rather, there is a need for developing guidelines, protocols and frameworks to inform development and refinement of such tools to achieve consistent and accurate responses to caregivers and patients of pediatric oncology.

From a clinician perspective, by automating the provision of detailed and understandable medical information, LLMs can reduce the time spent explaining common queries to patients and caregivers ^47^. In addition, LLM tools support the repetition of essential information at a time a caregiver chooses and as often as needed. However, LLMs have data cut-off points indicating the latest information LLMs have access to in their training data. Therefore, new evidence released after their cut-off point may not be available to include in their responses. This requires an additional disclaimer or real-time web access to check the latest information. The adaptability of LLMs to provide content based on specific patient contexts, as noted in the varying performance of different tools, underscores the potential for highly personalized patient education. This could lead to better understanding and retention of information; particularly as educational content, LLM tools could be adjusted based on the caregiver’s existing knowledge and informational needs.

### Study Limitations and Future Directions

This study has several limitations. The evaluation was based on a predefined set of questions derived from the COG Family Handbook and expert input, which may only cover the part of the full spectrum of caregiver inquiries. Additionally, the assessment process involved subjective evaluations by pediatric oncology experts (without patient or caregiver advocates), which could introduce bias. Moreover, the rapidly evolving nature of AI technology implies that the performance of LLMs may change with future updates, potentially affecting the generalizability and reproducibility of our findings over time. The concordance of experts in our study showed moderate agreement over LLM-tool performance, but the difference may hint at opposing interpretations of the evaluation criteria. Interrater reliability was similar between 5 experts compared to the literature ^48–50^. Evaluation criteria was not validated, instead it was extended from previous assessments of LLM-tools in the literature to meet the needs of the study methodology.

The study does not account for the dynamic interaction between caregivers and the LLMs. In real-world scenarios, caregivers might ask follow-up questions or seek clarifications and use prompt engineering, which was not simulated in this study. The static nature of our evaluation might only partially capture the conversational capabilities of the LLMs. Our study focused on publicly accessible LLM tools, which may not have the same capabilities as specialized or proprietary models used in clinical settings. The generalizability of our findings to other types of LLMs might be limited. The study’s cross-sectional design captures performance at a single point in time. We did not explore the integration of Retrieval-Augmented Generation (RAG), other knowledge base methods, or prompt engineering techniques ^51^, which may influence response quality. We aimed to evaluate these tools as laypersons typically use them, similar to a search engine, ensuring that the findings are relevant to real-world scenarios where users may need more specialized knowledge in prompt engineering or access to advanced retrieval methods.

Longitudinal studies are needed to understand how using LLMs impacts caregivers over time and how caregivers’ needs and LLM performance might evolve. In addition, future research should explore applying LLM tools in other medical specialties and patient demographics to assess their broader applicability. Integrating real-time feedback mechanisms into LLMs could enhance their adaptive learning capabilities with personalization, thereby improving the accuracy and contextual relevance of the information they provide.

## Conclusion

This study contributes to the growing body of literature on the applications of AI in healthcare by providing a detailed evaluation of how publicly available and prominent LLM-supported tools perform in a critical and sensitive domain. By identifying the strengths and limitations of these tools, this research offers insights about their potential role in enhancing caregivers’ information access and education in pediatric oncology. The findings can inform future developments in AI technology, aiming to improve the accessibility and quality of healthcare information for caregivers.

## Data Availability

All data produced in the present study are available upon reasonable request to the authors

**Appendix A.** Themes from Expert Evaluation Criteria Notes.

|  | Bard | Google SGE | Bing Chat | ChatGPT |
|---|---|---|---|---|
| **Accessibility of Language** | Many comments highlight that the language used is **too advanced** for the general public, including terms like "inherited mutations" and "lumbar puncture." There is a **need to simplify complex topics** like genetic mutations and biopsy steps using more accessible terms or even visual aids. | Assessors’ comments suggest that word choices **should be simplified** for better health literacy. One comment stated, "From a health literacy perspective, the word count is better; however, I still think the word choices could be simplified." | Some responses are described at **a grade level higher than recommended** for plain language, making it difficult for patients and families to understand. As one doctor noted, "This level of detail might be better shared in a conversation with providers." | Responses were **generally clear and well-organized**, with some being praised for hitting all major points and being easy to read. However, some responses used **advanced language and complex explanations** that would be difficult for non-medical consumers to understand without a healthcare professional's assistance. "Advanced language and explanations." "OMG. Where do I start? WAAAAY too complex" |
| **Accuracy of Information** | Assessors’ comments point out some **inaccuracies**, such as the incorrect information about urine tests detecting certain types of cancers or the inheritance of Down Syndrome.<br>Some responses are noted to be **too focused on adult patients** when the context is pediatric cancer. There's a **need for tailored information** specific to the patient group. | **Several inaccuracies** were noted, such as "one does not inherit cancer" and "Patients who are receiving cancer medications pose no risk to children, pregnant women, or anyone else." One doctor stated, "Statement 'one does not inherit cancer' is inaccurate for some rare forms of childhood cancer." Some statements are **misleading or incorrect**, such as "The Dana-Farber Cancer Institute offers a back-to-school program" which may not apply universally. "The statement that 'most children with cancer will attend school at least some of the time...' may not be true for all." | Some responses contain **inaccuracies or irrelevant information**. For instance, "The statistics provided about cancer incidence and mortality are not related to the question asked." **Incorrect** examples, such as mentioning "prostate" in the context of pediatric oncology, were highlighted: "Inclusion of 'prostate' as an example seems inappropriate." Responses sometimes provide **misleading** information, such as, "The answer goes beyond treatment options," | Several responses contained **inaccurate or incomplete information**. "Genetic factors are poorly explained and leaves room for misinterpretation. Including prenatal exposure to diagnostic x-rays also leaves room for parents to blame themselves and has a lack of evidence." Other responses were praised for being **accurate and comprehensive**. "Comprehensive, clear, well organized, accurate." "Concise - goes through all causes. Accurate." |
| **Completeness** | There were **mixed views on the completeness** of the information provided. Several comments acknowledge that the responses are thorough and cover a wide range of relevant details: "Very well done, clearly addressing the issue that there is not one definitive answer to this question". Some comments point out missing elements essential for a complete response. For example, "Information regarding the fact that the hair will likely re-grow after treatment is missing". There seems also more than the needed information: "This offers too much detail and offers too much additional information. It would have been better if the answer focused only on signs and symptoms of anemia." | Many responses are criticized for being **too vague or incomplete**. One comment noted, "Very limited information provided, overly broad." Some responses include **extraneous information** that can confuse readers. An assessor commented, "There is added information that is unhelpful to the reader and could add confusion." | Many responses **lacked crucial details** or only **partially answered** the question. One quote highlight this: "While it gives some of the answer, it needs more specifics to provide proper information." There were sometimes **missing key details**, like staging, treatment specifics, and follow-up care. For example, "None of the choices have alluded to the need for staging," and "Does not discuss follow-up appointments and what might be done." | Many responses were praised for their **thoroughness and specificity**, providing comprehensive information. Comments included, "Thorough and provides all the information" and "Very thorough and specific. Answer the question well." Despite overall thoroughness, some responses were **missing critical information** or needed more detail. "Response does not include information about anesthesia; use of the term "modern medicine" may be offensive to some groups. "Immunotherapy is missing from Leukemia." |
| **Practical Guidance** | Practical advice was sometimes missing. One doctor advised, "probably better to make sure the reader is aware that there are multiple possible explanations," suggesting that **practical context was needed**. The responses sometimes **failed to provide practical relevance**. One comment stated, "Need to expand on some things. Just saying risk factors exist is not enough." | Responses **lack practical advice** and clear instructions. For example, "This is too vague. Talking about spleen removal for anemia without talking about why you would do that could be frightening and confuse people." Some practical advice provided is sometimes **inapplicable**. For instance, "Keep tablets whole" is not always true and can be misleading. Some responses provide practical information that may be **inappropriate** for the context. For example, "We RARELY use hormone treatment in treating children with cancer," | Some information provided was **impractical or irrelevant** to the specific context. For example, "Doesn't seem completely related to cancer. Not all answer are practical,". **Unrealistic or impractical suggestions** reduced the utility of some responses: "Doesn't seem completely related to cancer. Not all answer are practical, i.e. 'find other things to do with hands' and 'get professional help'." | **Practical suggestions** and options were appreciated. "Good lists of options, with nice to have examples", "Good answer - gives specific things that can be done. Liked the part of when to seek immediate medical care. Other answers did not discuss this" Some responses were seen as **too generic or not tailored enough** for the specific situation. "Contains some accurate information that is clearly conveyed but it is not tailored for a child receiving cancer treatment."<br>"Accurate but generic" |
| **Emotional Tone and Empathy** | Mixed reactions to the empathetic tone in responses, with some finding it inappropriate or too personal coming from a computer. While some comments appreciate the attempt to provide emotional support, others indicate it may be **misplaced or overly personal**. "While I like the empathy this response attempted, I didn't respond well to the level of intimacy if this is coming from a bot." | Some responses **fail to provide the necessary emotional support** and reassurance that patients and their families need. For instance, assessors noted that responses were often "too medical/technical" when the questions seemed to be seeking emotional support rather than just medical facts. One comment mentioned, "Too medical/technical - the question appears to be asking for emotional support, not medical support." Furthermore, phrases like "Doctors cut out tissue" were criticized for being **too blunt**. | The use of supportive and empathetic language was occasionally seen as **unnecessary or misdirected**. For example, "I hope this helps. Let me know if you have any other questions," was seen as extraneous. The tone of some responses was critiqued **for lacking empathy or being inappropriate**. For instance, "People do not want to hear this, 'Treatment may result in remission, recurrence, or no response.'" and "The empathetic statement at the beginning of the text is unnecessary."<br>Avoiding harsh tones was viewed as important: "Incomplete and the last section has a punitive tone." | **Empathetic tones** were appreciated for some prompts.<br>"Good emotional support and practical things to help"<br>Clinical tones without empathy were noted for other prompts.<br>"The empathetic statement at the beginning of the text is unnecessary." There was a noted **lack of cultural considerations** in some responses, with one critique stating, "Information regarding different cultural considerations is missing." |

